# Prenatal *BRCA1* epimutations contribute significantly to triple-negative breast cancer development

**DOI:** 10.1101/2023.05.14.23289949

**Authors:** Oleksii Nikolaienko, Hans P. Eikesdal, Bjørnar Gilje, Steinar Lundgren, Egil S. Blix, Helge Espelid, Jürgen Geisler, Stephanie Geisler, Emiel A.M. Janssen, Synnøve Yndestad, Laura Minsaas, Beryl Leirvaag, Reidun Lillestøl, Stian Knappskog, Per E. Lønning

## Abstract

**Background:** Normal cell *BRCA1* epimutations have been associated with increased risk of triple-negative breast cancer (TNBC). However, the fraction of TNBCs that may have *BRCA1* epimutations as their underlying cause is unknown.

**Methods:** To address this question, we analyzed *BRCA1* methylation status in breast cancer tissue and matched white blood cells (WBC) from 411 patients with primary breast cancer, including 66 TNBCs, applying a highly sensitive sequencing assay, allowing allele-resolved methylation assessment. Further, to assess the time of origin and the characteristics of normal cell *BRCA1* methylation, we analyzed umbilical cord blood of 1260 newborn girls.

**Results:** We found concordant tumor and mosaic WBC *BRCA1* epimutations in 10 out of 66 patients with TNBC and in four out of six patients with estrogen receptor (ER)-low expression (<10%) tumors (combined: 14 out of 72; 19.4%; 95% CI 11.1–30.5). In contrast, we found concordance in only three out of 221 patients with ER≥10% tumors and zero out of 116 patients with HER2-positive tumors. Intraindividually, *BRCA1* epimutations affected the same allele in normal and tumor cells. Assessing *BRCA1* methylation in umbilical WBCs from girls, we found mosaic, predominantly monoallelic *BRCA1* epimutations, with qualitative features similar to those in adults, in 113/1260 (9.0%) of individuals.

**Conclusions:** Our findings reveal prenatal *BRCA1* epimutations to be the underlying cause of around 20% of TNBC and low-ER expression breast cancers.

## Background

Aberrant gene promoter methylation, or epimutations, are observed in many cancer types. While such epimutations may be passenger events of limited biological importance, it is well established that promoter methylation of tumor suppressor genes (TSGs) may contribute as driving forces in tumor progression [1, 2].

*BRCA1* germline mutations are the most frequent cause of hereditary breast and ovarian cancers [3]. Most cancers arising in *BRCA1* mutation carriers belong to the triple-negative subclass of breast cancers (TNBC) and the high-grade serous subclass of ovarian cancers (HGSOC). Contrasting *BRCA2* [4], *BRCA1* is frequently methylated in sporadic TNBC and HGSOC tumors [5–7], and it is well established that such promoter methylation is associated with repressed *BRCA1* transcription [8, 9]. TNBCs with *BRCA1* methylation have a gene expression profile closely resembling TNBCs arising in *BRCA1* mutation carriers [5]. While *BRCA1* promoter methylation and *BRCA1* mutations seem to a large extent to be mutually exclusive in both TNBCs and HGSOCs [5, 10], conflicting evidence indicates similarities and differences between *BRCA1* promoter methylation and mutations regarding therapy sensitivity in breast cancer [6, 11, 12].

Constitutional epimutations are defined as aberrant normal tissue methylation occurring in early life, generally affecting all three germ layers [13]. There are two types; secondary epimutations, caused by specific genetic aberrations, and primary epimutations, for which no underlying genetic factor is found [13]. Contrasting secondary epimutations, primary epimutations often present in a low-level, mosaic pattern, affecting only a small fraction of cells [10]. While secondary constitutional methylation of *BRCA1* has been observed in a few families with an elevated risk of breast and ovarian cancer [14–16], the question of primary constitutional methylation as a cancer risk factor has remained controversial [4, 10, 17-23]. However, in a recent study we found white blood cell (WBC) *BRCA1* promoter methylation to predict an elevated risk of incident TNBC as well as HGSOC >5 years after blood sampling in healthy women [24]. While these findings indicate that *BRCA1* methylation may arise in normal cells subsequently developing into cancer precursors, several key questions need to be addressed. First, we do not know whether WBC *BRCA1* mosaic methylation arise prenatally (constitutional) or may be acquired postnatally (somatic normal tissue methylation). In case of constitutional methylation, we need to address whether such methylation may be fully developed across the promoter prenatally or exist as a precursor for subsequent development at a later stage. Second, in case *BRCA1* methylated cells arising by a prenatal clonal expansion as constitutional methylation, one would expect a qualitatively similar, allele-specific WBC methylation in newborns as that recorded in adults. Third, if *BRCA1*-methylated WBCs represent constitutional methylation, and methylated WBCs and breast cancer precursor cells share a common embryonic clonal origin, one would expect a similar allele-specific *BRCA1* methylation profile [24] in WBCs and matched *BRCA1*-methylated tumors from the same individual. Fourth, we need to assess the fraction of TNBCs arising from constitutionally *BRCA1*-methylated cells, i.e. the fraction of TNBCs, previously considered as “sporadic”, that could be explained by underlying *BRCA1* methylation. While we recently reported the hazard ratio for incident TNBC with respect to WBC *BRCA1* methylation to be 2.35 [24], such a hazard ratio provides an indirect estimate for the fraction of tumors actually derived by this mechanism [25]. Moreover, the fact that the median age of women enrolled in our previous study was 62 years, indicates that a substantial fraction of TNBCs that may have been overlooked due to diagnosis at a younger age.

To address these questions, we evaluated the incidence, magnitude, intraindividual tissue concordance and allele specificity of *BRCA1* methylation in tumor and matched WBC from 411 patients with primary breast cancer, including 66 TNBCs. In addition, we analyzed WBC *BRCA1* methylation in 1260 umbilical cord blood samples from newborn girls.

## METHODS

### Patients and tissue sampling

In the present study, we included all patients enrolled in three neoadjuvant breast cancer studies (EPITAX, DDP and PETREMAC) [6, 26, 27] from which pretreatment tumor tissue and WBC DNA samples were available for analysis (Fig. 1). The studies were approved by the Regional Ethics Committee (273/96-82.96, 06/3077 and 2015/1493), and all patients provided written informed consent at enrolment. The DDP and PETREMAC trials were registered under ClinicalTrials.gov (NCT00496795 and NCT02624973) while the EPITAX was conducted prior to ClinicalTrials implementation.

**Fig. 1.**
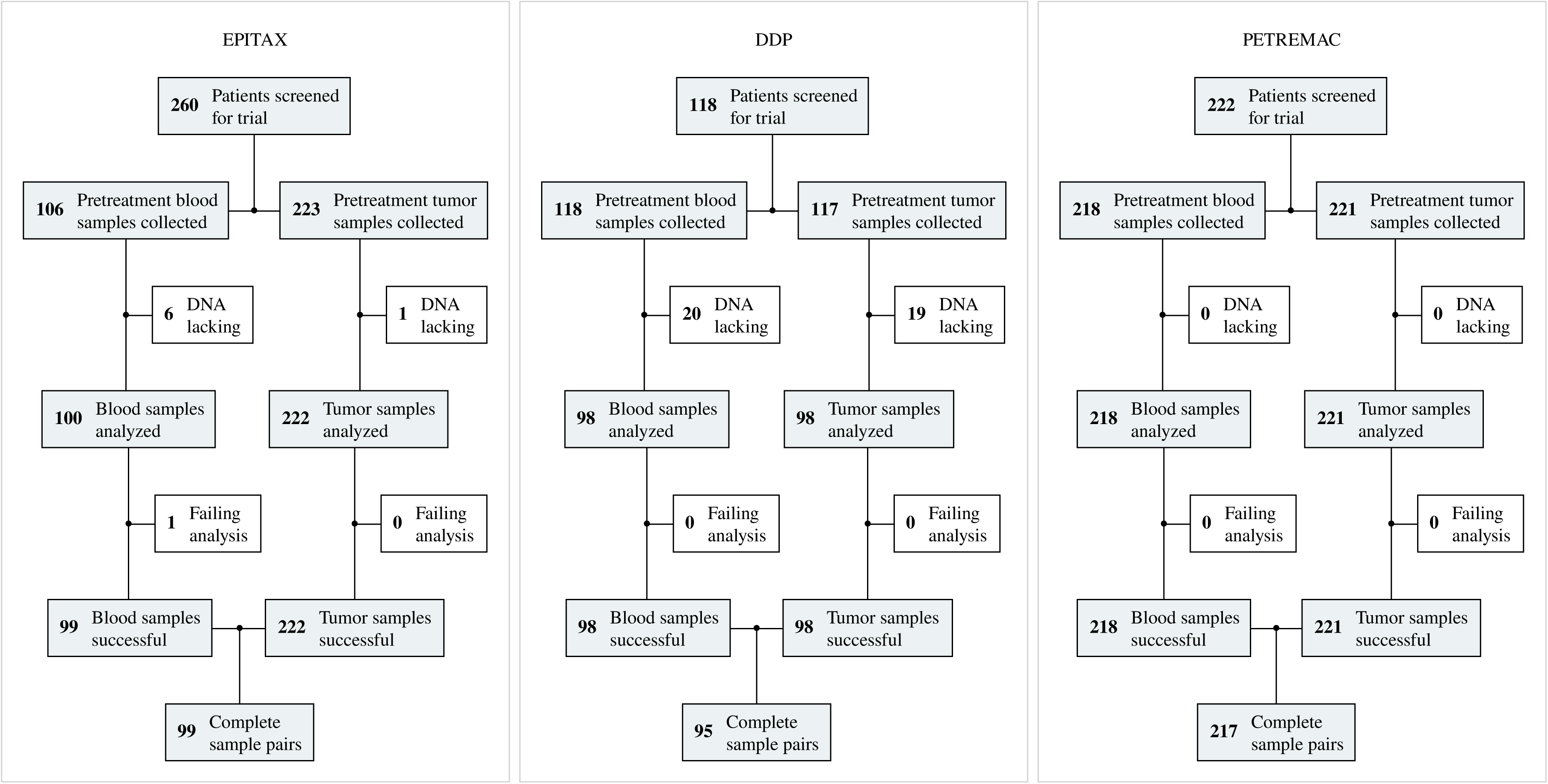
CONSORT diagram depicting patient enrolment in the EPITAX, DDP and PETREMAC clinical trials, and the number of pretreatment samples collected and successfully analyzed in the current study.

All patients underwent an incisional or Tru-Cut tumor biopsy prior to treatment. Tumor biopsies were snap-frozen in liquid nitrogen at removal and stored in liquid nitrogen, while WBC specimens were stored at –80°C after centrifugation of EDTA whole blood and plasma removal.

The Norwegian Mother, Father and Child Cohort study (MoBa) is an ongoing cohort study enrolling more than 110 000 newborns and their parents [28]. For the present study, we randomly selected 1260 newborn girls, including 420 born prematurely (before 36 weeks of gestation) and 840 girls born at normal term (39–41 weeks of gestation). Procedures for sample collection, DNA extraction and storage have been described previously [29]. No difference in *BRCA1* methylation was observed between premature and normal term newborns, and the samples were therefore treated as a unified cohort in the present study.

### Sample preparation

Procedures for DNA and RNA extraction from tumor and WBC samples are outlined in Additional file 1. In brief, genomic DNA for methylation analyses was extracted from tumor and WBC samples using QIAamp DNA Mini kit (Qiagen, Valencia, CA) and total RNA for gene expression analysis was extracted from tumor tissue using the RNeasy Mini kit (Qiagen, Valencia, CA).

### Methylation sequencing

For *BRCA1* methylation analysis, DNA bisulfite conversion, amplification and sequencing was performed as described previously [24] Briefly, 500 ng of genomic DNA were bisulfite converted and subjected to *BRCA1* gene promoter fragment amplification using four pairs of primers, that do not overlap with any of the CpG dinucleotides (GRCh38 genomic coordinates: CpG00–13 chr17:43125624–43126026, CpG14–31 chr17:43125270–43125640, CpG17–34 chr17:43125171–43125550, CpG33–49 chr17:43124861–43125249; Supplementary Fig. S1). All four amplicons were combined, indexed, and sequenced by 2×226 bp reads using Illumina MiSeq System (Illumina, San Diego, CA), resulting in an ultradeep coverage of about 30000x for each amplicon. As previously described for case-control analyses [24], the region covering CpGs 17–34 is considered biologically relevant and was used as the main measure for methylation calling. The overlapping amplicon covering CpGs 33–49, also covered SNP rs799905, and was used for allele-specific methylation assessment.

### Molecular subtyping

Estrogen and progesterone receptor (ER, PgR) as well as HER2 status was determined upon inclusion in each of the clinical trials. For the present analyses, we defined the cutoff for ER positivity as 1%. Since the EPITAX trial, conducted in 1997–2003, used a cutoff of 10%, all cases where ER status was recorded as <10% were re-examined according to standard criteria and classified as either <1% or 1–9%.

All tumors were assigned to intrinsic subtypes based on mRNA expression profiling according to the classification by Perou *et al.* [30] using either RNA sequencing (DDP and PETREMAC trials) or mRNA microarrays (EPITAX trial) (for details, see Additional file 1). While the percentage of ER-positive tumors is lower and the percentage of HER2+ and TNBC/ER-low tumors is higher in the oldest (EPITAX trial) subset, as compared to the DDP and PETREMAC trials, the same difference is present in the full study population, [6, 26, 27] reflecting a higher enrolment of patients with HER2+/TNBC when the EPITAX trial was actively recruiting in the 1990s.

### *BRCA1* mutation status assessment

Data on *BRCA1* mutation status for patients were collected from our previous genetic analyses [6, 27]. For cases lacking previous genetic data, we performed targeted sequencing of a cancer gene panel, as previously described, [31] and drew *BRCA1* mutation status from the generated data. For consistency, all detected variants were re-audited for pathogenicity according to the ClinVar database [32].

### Data analyses

For *BRCA1* methylation analysis, sequencing reads were mapped/aligned to the GRCh38 reference genome using the Illumina DRAGEN Bio-IT Platform (v3.6.3). Cytosine methylation and its allele specificity was evaluated using the epialleleR R package (v1.3.5) [33]. A single quantitative metric of methylation (hypermethylated variant epiallele frequency, VEF) was obtained by averaging frequencies of hypermethylated epialleles for two amplicons covering CpGs 14–31 and 17–34 as previously reported [24]. The cutoffs for methylation positivity were determined computationally, following the same predefined approach as previously reported [24] (for details, see Additional file 1). The cutoffs were defined as 6.96×10^−4^ for *BRCA1* methylation in WBC and 4.71×10^−2^ for tumors. The differences between the cutoffs in WBC and tumors also reflect a biological rationale: WBC *BRCA1* methylation is expected to present a low-level, mosaic pattern [10, 34], while tumors arising from *BRCA1*-methylated cells are expected to harbor a large fraction of methylated cells due to clonal expansion. Notably, for sensitivity analyses all WBC data from newborns in the present study were also re-assessed applying the cutoff defined by WBC-analyses of cancer patients and vice versa. These sensitivity analyses had no impact on the biological conclusion (see Additional file 1 for details).

### Statistical analysis

Correlation in methylation status between tumor and corresponding WBC or normal breast tissue samples were evaluated using Fisher’s exact test. Methylation frequencies were presented with confidence intervals. R software environment for statistical computing (v4.1.2) was used for all statistical analyses.

## RESULTS

### Study population characteristics

To assess *BRCA1* methylation in breast cancer patients, we included all patients enrolled in three neoadjuvant breast cancer studies (the EPITAX, DDP and PETREMAC trials [6, 26, 27] from whom pretreatment tumor tissue and matched WBC DNA samples were available. Among a total of 600 patients screened, 411 had both blood and tumor pretreatment samples successfully analyzed and were included in the final results. The reasons for exclusion are presented in Fig. 1 and details of the patients included are given in Table 1.

**Table 1.** Patient characteristics in the EPITAX, DDP and PETREMAC clinical trials. TNBC = triple-negative breast cancer, HER2 = Human epithelial-like receptor-2, ER = estrogen receptor, basal = basal-like gene expression profile, Her2 = HER2-enriched gene expression profile, LumA = luminal A gene expression profile, LumB = luminal B gene expression profile, Normal = normal-like gene expression profile, IDC = invasive ductal carcinoma, ILC = invasive lobular carcinoma.

|  | EPITAX<br>(N = 99) | DDP<br>(N = 95) | PETREMAC<br>(N = 217) |
| --- | --- | --- | --- |
| **Age** |  |  |  |
| Mean (SD) | 49.5 (10.3) | 48.4 (9.80) | 53.0 (10.9) |
| Median [Min, Max] | 49.0 [25.0, 70.0] | 48.0 [24.0, 71.0] | 51.0 [27.0, 78.0] |
| **Tumor receptor status** |  |  |  |
| TNBC | 16 (16.2%) | 17 (17.9%) | 33 (15.2%) |
| HER2-/ER<10% | 4 (4.0%) | 0 | 2 (0.9%) |
| HER2-/ER≥10% | 48 (48.5%) | 59 (62.1%) | 114 (52.5%) |
| HER2+ | 29 (29.3%) | 19 (20.0%) | 68 (31.3%) |
| Missing | 2 (2.0%) | 0 | 0 |
| **Tumor molecular subtype** |  |  |  |
| Basal | 24 (24.2%) | 14 (14.7%) | 29 (13.4%) |
| Her2 | 20 (20.2%) | 14 (14.7%) | 45 (20.7%) |
| LumA | 22 (22.2%) | 32 (33.7%) | 70 (32.3%) |
| LumB | 22 (22.2%) | 25 (26.3%) | 51 (23.5%) |
| Normal | 11 (11.1%) | 10 (10.5%) | 20 (9.2%) |
| Missing | 0 | 0 | 2 (0.9%) |
| **Tumor histology** |  |  |  |
| IDC | 80 (80.8%) | 72 (75.8%) | 159 (73.3%) |
| ILC | 15 (15.2%) | 17 (17.9%) | 32 (14.7%) |
| Other | 4 (4.0%) | 6 (6.3%) | 26 (12.0%) |

To assess allele-specific mosaic *BRCA1* methylation in newborns, 1260 umbilical cord blood samples were drawn from the Norwegian Mother, Father and Child Cohort study (MoBa) [35], as listed in Table 2.

**Table 2.**
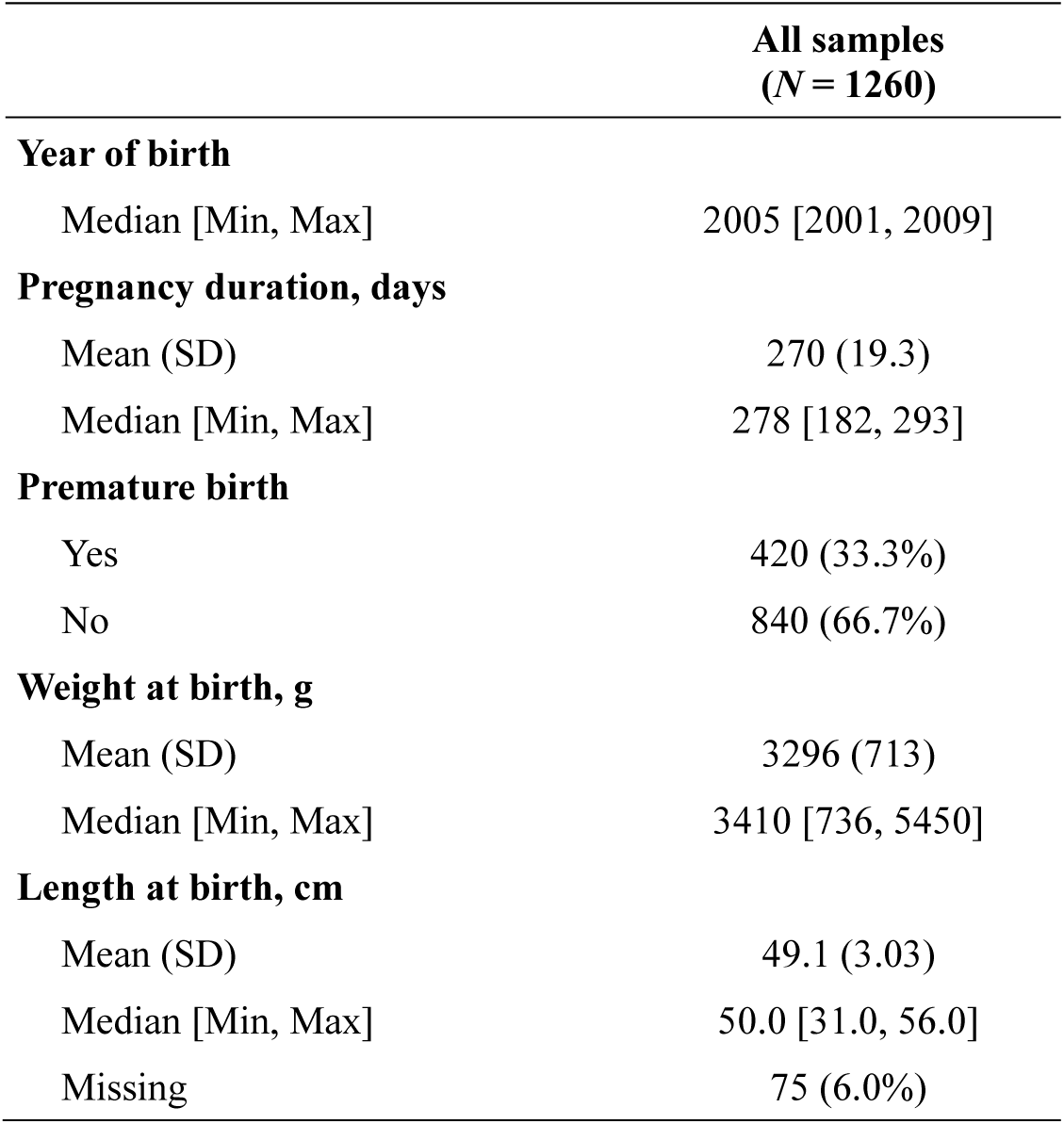
Participant characteristics of newborns drawn from the MoBa trial.

|  | All samples<br>(*N* = 1260) |
| --- | --- |
| **Year of birth** |  |
| Median [Min, Max] | 2005 [2001, 2009] |
| **Pregnancy duration, days** |  |
| Mean (SD) | 270 (19.3) |
| Median [Min, Max] | 278 [182, 293] |
| **Premature birth** |  |
| Yes | 420 (33.3%) |
| No | 840 (66.7%) |
| **Weight at birth, g** |  |
| Mean (SD) | 3296 (713) |
| Median [Min, Max] | 3410 [736, 5450] |
| **Length at birth, cm** |  |
| Mean (SD) | 49.1 (3.03) |
| Median [Min, Max] | 50.0 [31.0, 56.0] |
| Missing | 75 (6.0%) |

### Concordant WBC and tumor *BRCA1* methylation

Based on the previously detected association between constitutional *BRCA1* methylation and risk of TNBC, we analyzed concordance of *BRCA1* methylation in tumors and matched WBC samples to assess the fraction of TNBCs potentially caused by underlying constitutional *BRCA1* methylation. In total, 17 out of 66 (25.8%; 95% CI 15.8–38.0) patients with TNBC harbored tumor *BRCA1* methylation (Table 3). Notably, among these 17 patients, 10 (58.9%; CI 32.9–81.6%) also carried WBC *BRCA1* methylation (WBC and tumor tissue methylation concordance: *P* < 0.001; Fig. 2). Thus, 15.2% (95% CI 7.5–26.1%) of all TNBCs revealed concordant tumor and WBC *BRCA1* methylation.

**Fig. 2.**
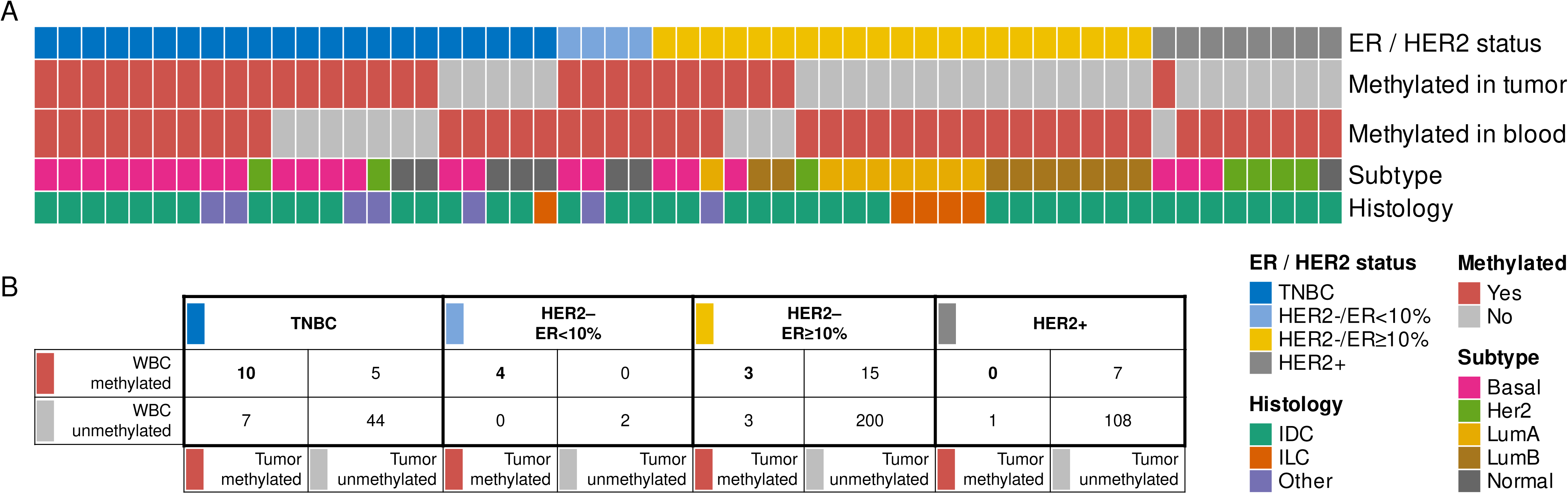
*BRCA1* methylation in matched blood and tumor samples in breast cancer patients. (**A**) Molecular and histological characteristics (rows) of all samples (*N* = 55; columns) belonging to matched sample pairs carrying *BRCA1* methylation in the blood (WBC) and/or tumor. TNBC = triple-negative breast cancer, HER2 = Human epithelial-like receptor-2, ER = estrogen receptor, basal = basal-like gene expression profile, Her2 = HER2-enriched gene expression profile, LumA = luminal A gene expression profile, LumB = luminal B gene expression profile, Normal = normal-like gene expression profile, IDC = invasive ductal carcinoma, ILC = invasive lobular carcinoma. (**B**) Concordance of *BRCA1*-methylation status in WBC and tumor tissue among all patients analyzed, stratified for tumors belonging to the different breast cancer subgroups.

Regarding tumors with ER expression within 1–9%, genomic profiling has revealed these tumors to mirror gene expression profiles recorded in TNBC [36]. In this subgroup, four out of six patients harbored *BRCA1* methylation in their tumor tissue, all revealing concordant WBC methylation (Fig. 2). Grouping the TNBC and ER-low (1–9%) tumors together, concordant tumor and WBC *BRCA1* methylation was observed in 14 out of 72 patients (19.1%; 95% CI 11.1–30.5%). Notably, these 14 constituted the majority of the 21 patients with *BRCA1* methylated TNBC or ER low (1–9%) tumors (66.7%; 95% CI 43.0–85.4%).

For HER2-negative tumors expressing ER≥10%, six out of 221 revealed *BRCA1* tumor methylation with only three of these patients revealing concordant WBC *BRCA1* methylation. Further, the lowest methylation frequency was observed among HER2-positive tumors (independent of ER expression). Here, one out of 116 tumors revealed *BRCA1* methylation in the tumor tissue, and this patient was negative for WBC *BRCA1* methylation (Fig. 2).

As for patients harboring *BRCA1* methylation in both tumor and WBC, *BRCA1* methylation levels in the tumors were 36–103 fold higher than in blood, consistent with clonal expansion of cells with methylated *BRCA1* alleles (Fig. 3A). Intratumoral levels of *BRCA1* methylation were similar for TNBCs (7.6–78.8%), ER-low (10.7–36.4%), and the remaining non-TNBC methylated tumors (13.8–82.4%).

**Fig. 3.**
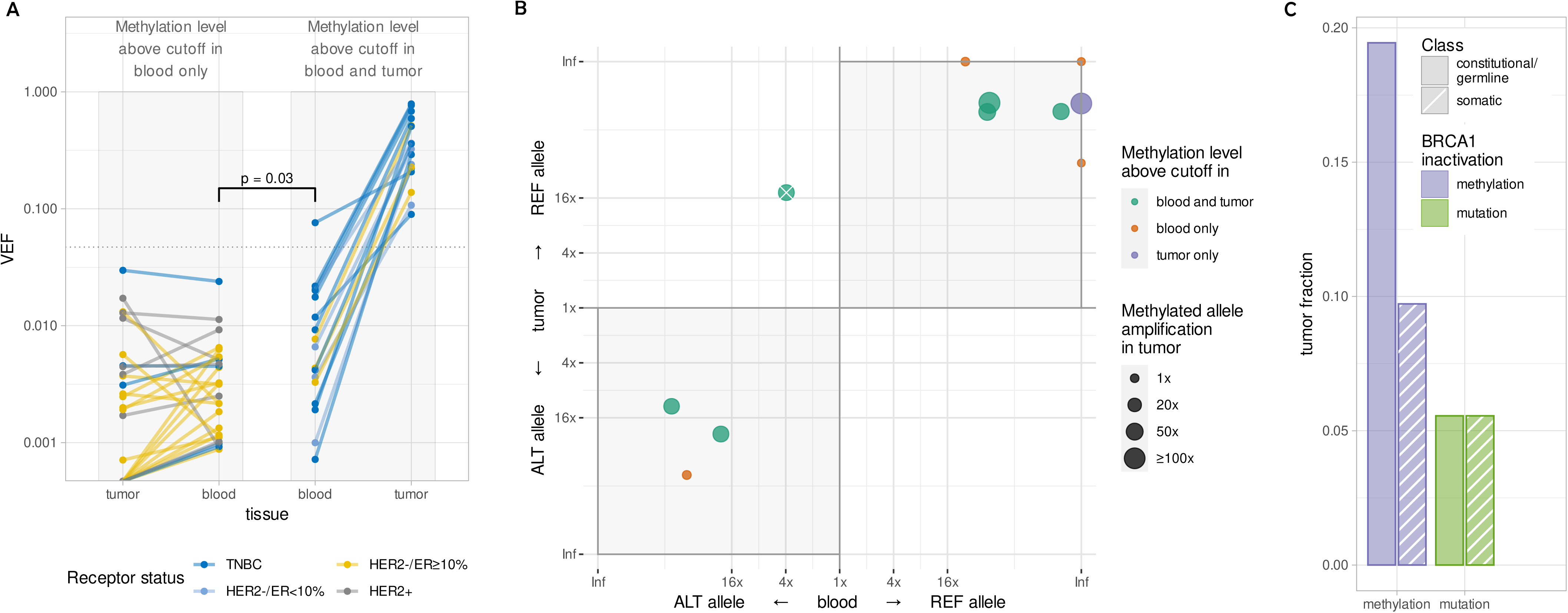
(A) Quantitative levels of *BRCA1* methylation (VEF value for region CpG14–34) in blood and tumor samples of breast cancer patients from whom blood samples had *BRCA1* methylation levels above the blood-specific cutoff. Solid lines connect matched samples; lines and dots are colored according to tumor receptor status. Gray boxes outline patients with *BRCA1* methylation *not enriched* (left) or *enriched* (right) in tumors; dotted line represents cutoff value for *BRCA1* methylation positivity in tumor tissue. Both quantitative (ANOVA) and qualitative (Wilcoxon rank sum) tests confirm significant difference between subsets of blood VEF values (shown by square bracket). (**B**) Allele specificity of *BRCA1* methylation in blood and tumor samples from breast cancer patients heterozygous for SNP rs799905 (*N* = 11). Preferential methylation of one of the alleles is evaluated and plotted as fold enrichment, with allele-specific preference in methylation in blood on the x-axis and in tumor on the y-axis. Gray shading indicates quadrants supporting concordant allelic methylation in matched blood and tumor. Data points falling in the upper-right quadrant indicate the reference allele of rs799905 to be the predominantly methylated allele in blood and tumor, while data points falling in the lower-left quadrant indicate the alternative allele of rs799905 to be the predominantly methylated allele in blood and tumor. Dots (representing matched sample pairs) are colored according to *BRCA1* methylation status in tumor and blood with their size representing fold amplification of the methylated allele in tumor tissue compared to the corresponding blood sample. The crossed-out dot represents an individual with comparable methylation of both alleles in blood but predominantly the reference-allele methylated in tumor, likely reflecting the tumor to have originated from one out of two methylated lineages of normal cells (see main text). Inf, infinity value, i.e., exclusive methylation of a single allele. (**C**) Fractions of TNBC and HER2–/ER<10% tumors (*N* = 72) characterized by different molecular mechanisms of *BRCA1* inactivation (methylation [blue] or mutation [green]) and its potential time of emergence (constitutional/germline [solid fill] or somatic [stripe pattern]).

Taken together, these findings indicate that around 20% of all TNBC/ER-low breast cancers and around 65% of all *BRCA1*-methylated tumors occur in individuals with underlying constitutional *BRCA1* methylation.

### Allele-specific concordance of *BRCA1* methylation in WBC and tumor tissue

While concordant *BRCA1* methylation in tumor tissue and matched blood samples may suggest a common clonal origin, we sought to provide further evidence for this hypothesis by assessing the allele specificity of the *BRCA1* methylation in tumors and blood.

Allele-specific methylation may be detected in cases heterozygous for the SNP rs799905, since this polymorphism is located in the area that is covered by the *BRCA1* methylation assay (see Methods and Additional file 1; Fig. S1). Among 17 patients carrying concordant *BRCA1* methylation in WBC and tumor tissue, SNP rs799905 genotype information was lacking and/or could not be linked to methylation in two patients. For the 15 informative individuals, seven were homozygous for the reference allele, two were homozygous for the alternative allele, while six were heterozygous. The allelic distribution of *BRCA1* methylation for WBCs and tumor samples among these six informative heterozygous cases is depicted in Fig. 3B (green dots). *BRCA1* methylation was enriched on the same allele in the tumor tissue and WBC in five of these individuals, indicating a shared clonal origin of the methylated normal and tumor cells. The sixth patient revealed comparable levels of *BRCA1* methylation of both rs799905 alleles in blood (with a slight preference for methylation on the alternative allele) while the tumor carried methylation of the reference allele. Most likely, this patient harbored two independent subclones of *BRCA1*-methylated normal cells, with one clone giving rise to the tumor cells.

As the tumor samples were not subject to microdissection, they contain a number of different types of benign cells including normal breast epithelium, fibroblasts, circulating WBCs and macrophages [37]. Among individuals harboring WBC but not tumor *BRCA1* methylation (n = 27), 17 revealed small traces of *BRCA1*-methylated cells in the tumor biopsies, below the defined threshold for classification of tumors as methylation-positive but above the methylation threshold applied to WBC samples. This is consistent with low-level mosaic *BRCA1* methylation in normal breast cells and/or other normal cells present in the tumor biopsies. Among these 17 patients, four were heterozygous for rs799905 and thus informative for allele-specific methylation status. These four all revealed the low-level *BRCA1* methylation in their tumor biopsies to share the same magnitude and allele specificity as the methylation in the matched WBCs (Fig. 3B, red dots).

In addition, one patient with a *BRCA1* methylated TNBC and WBC *BRCA1* methylation just below the formal cutoff for positivity could be assessed for allelic methylation concordance. This patient also revealed a similar allele specific *BRCA1* methylation in tumor and WBC samples (Fig. 3B, blue dot).

Taken together, these findings reveal an allelic concordance between *BRCA1* methylation in WBC and matched cancer or benign tissue in the breast cancer samples, indicating that the methylated tumors have arisen from methylated normal cells in the affected mosaic individuals.

### Intrinsic breast cancer subtypes and *BRCA1* methylation

The distribution of *BRCA1* methylation was determined among the intrinsic subtypes of breast cancers, based on their mRNA signatures [30]. The subtype distribution of *BRCA1-*methylated tumors did not differ between TNBCs and non-TNBCs, neither was there a difference between those harboring concordant WBC *BRCA1* methylation and those that did not (Fig. 2A). Regarding TNBCs and ER<10% tumors with concordant tumor and WBC *BRCA1* methylation, 11 out of 14 were basal-like, two were normal-like, while one tumor expressed a HER2-enriched profile, despite absence of *HER2* gene amplification or positive protein staining. Interestingly, among the three ER≥10% tumors revealing concordant tumor and WBC *BRCA1* methylation, two revealed a basal-like profile, while the remaining one was classified as luminal A.

### *BRCA1* mutations and methylation

Among the 411 tumors analyzed, nine harbored *BRCA1* pathogenic variants (four somatic and five germline; Fig. 3C; Additional file 1; Table S1). None of the patients with germline *BRCA1* pathogenic variants revealed either WBC or tumor *BRCA1* methylation. While one patient with a somatic *BRCA1* mutation harbored tumor tissue *BRCA1* methylation in concert, no *BRCA1* methylation was detected in this patient’s WBCs, indicating the tumor *BRCA1* methylation, similar to the mutation, to be a somatic event.

### Frequency and allele specificity of *BRCA1* methylation in WBC from newborns

To assess the hypothesis of early origin (Fig. 4A) and potential dynamics of *BRCA1* epimutations and their allele specificity, we analyzed umbilical cord blood samples from 1260 newborn girls. We found 113 out of 1260 newborns to be *BRCA1* methylation-positive (9.0%; 95% CI 7.5–10.7%), with average beta value distribution (Fig. 4B) as well as hypermethylated variant epiallele frequencies (VEF) in a very similar range as seen in adult breast cancer patients (7.0×10^-4^ to 5.9×10^-2^ and 7.2×10^-4^ to 7.6×10^-2^, respectively; p>0.1; Fig. 4D). Also, similar to the intramolecular methylation pattern seen in adults, there was a relatively sharp distinction between non-methylated and hypermethylated epialleles, with the majority of methylation-positive epialleles being close to fully methylated (i.e. methylated on all CpGs), and very few epialleles having intermediate methylation levels (i.e. methylated at 20-80% of epialleles; Fig. 4C). No particular single CpG or stretch of consecutive CpGs in the methylated region stood out as systematically more or less methylated than others.

**Fig. 4.**
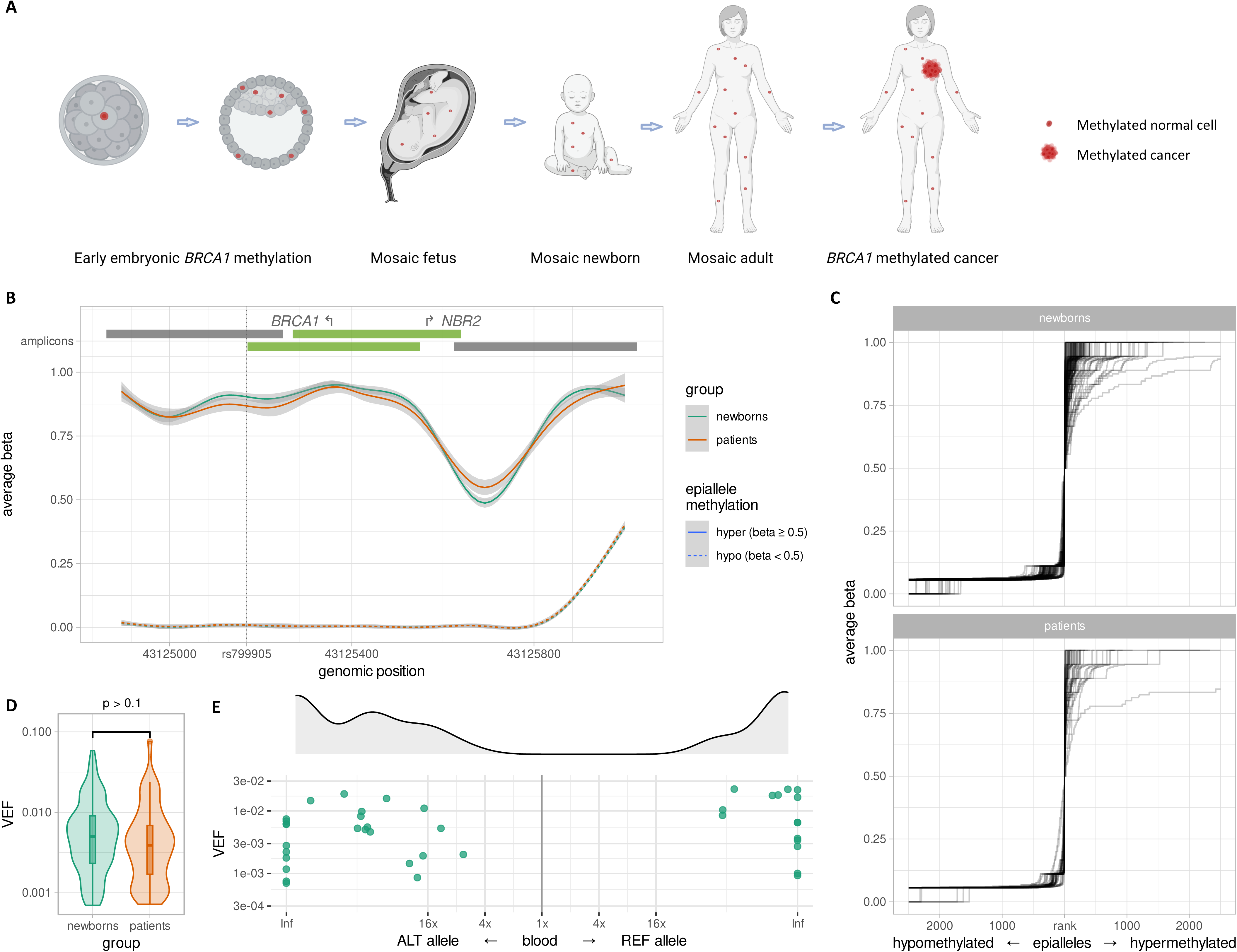
Similar properties of *BRCA1* methylation in blood samples of healthy newborn girls and adult breast cancer patients (**A**) Overall model for early prenatal (constitutional) *BRCA1* methylation as an underlying contributor to TNBC. Red dots represent *BRCA1* methylated normal cells, appearing through an early embryonic event, resulting in a mosaic adult. Red star represent breast cancer. (**B**) Smoothed averaged CpG methylation levels (y-axis) within assayed genomic region (x-axis) in blood of *BRCA1* methylation-positive newborn girls (*N*=113; green lines) and breast cancer patients (*N*=44; red lines). Solid lines represent averages for all hypermethylated epialleles (per-epiallele average beta value ≥ 0.5); dashed lines represent averages for all hypomethylated epialleles (per-epiallele average beta value < 0.5); light gray areas represent 95% CI. Bars on top represent amplicons, with the bright green ones covering CpGs 14–34. Arrows show *BRCA1* and *NBR2* transcription start sites; vertical dotted line marks position of SNP rs799905 (see Supplementary Information for more details). (**C**) Average beta values (y-axis) of ranked epialleles (x-axis) in blood samples of *BRCA1* methylation-positive newborn girls (*N* = 113; top) and breast cancer patients (*N* = 44; bottom). All epialleles of the region CpG14–34, within each sample, were ranked by increasing average beta value with every rank centered at epiallele with average beta value of 0.5. Lines connect increasing beta values and represent individual samples. Maximum 5000 epialleles are plotted per sample (beta = 0.5, +/-2500 alleles). The sharp incline in average beta value around beta = 0.5 reveals that most alleles are either hypomethylated or hypermethylated; very few alleles have intermediate methylation levels. (**D**) Density plot of *BRCA1* methylation levels (VEF value for region CpG14–34) in blood samples of newborn girls (*N* = 113; green) and breast cancer patients (*N* = 44; red). The lower and upper hinges of boxes correspond to the first (Q_1_) and third (Q_3_) quartiles, respectively; the bar in the middle correspond to the median value; the upper and lower whisker extend to Q_3_+1.5*IQR and Q_1_–1.5*IQR, respectively, while the values outside this range (outliers) are plotted as dots. Neither quantitative (ANOVA) nor qualitative (Wilcoxon rank sum) test show significant difference between these sets of blood VEF values (shown by square bracket). (**E**) Allele specificity of *BRCA1* methylation in blood samples from newborn girls heterozygous for SNP rs799905 (*N* = 40). Preferential methylation of one of the alleles (fold enrichment) is indicated on the x-axis and degree of methylation (VEF value for region CpG14–34) is indicated on the y-axis. Data points in the right half of the plot indicate methylated alleles to be predominantly rs799905 reference alleles, while data points towards the left indicate methylated alleles to be predominantly rs799905 alternative alleles. Gray area above the plot shows smooth kernel density estimates for fold enrichment values. Inf, infinity value, i.e., exclusive methylation of a single allele.

Among the 113 newborns revealing WBC *BRCA1* methylation, sufficient amounts of DNA for SNP rs799905 assessment were available from 89. Among these, we found 40/89 (44.9%) of the girls to be heterozygous for SNP rs799905, while 12/89 (13.5%) and 37/89 (41.6%) were homozygous for the alternative and reference alleles, respectively. This distribution is similar to that recently established in adult women in the US Women’s Health Initiative study [24]. Intraindividually, among the 40 girls heterozygous for rs799905, methylation was located predominantly on one specific allele, similar to the pattern seen in adults (Fig. 4E). Further, across these heterozygous individuals, there was a similar overall distribution of methylated alleles between the SNP rs799905 alternative and reference allele (p > 0.10).

Taken together, our findings confirm *BRCA1* epimutations in newborns to be predominantly monoallelic intraindividually but to occur independently of promoter haplotype across individuals, and with low VEF values, resembling the findings in adults.

## DISCUSSION

Epigenetic regulation plays a key role in normal cell function during life and is influenced by genetic as well as environmental factors. Epimutations may occur either as somatic events during life or as so-called constitutional methylation, arising *in utero*, in which case it may affect normal tissues derived from all three germ layers [13]. While epimutations causing gene silencing of tumor suppressors like *MGMT*, *MLH1* and *BRCA1* are well known across many malignancies [6, 8, 38-41], in general such epimutations are considered somatic. However, the seminal discovery by Hitchins and colleagues of inherited *MLH1* methylation in a family prone to colon cancer [42] sparked interest in constitutional methylation of TSG promoters as a potential underlying cause of cancer [13, 43].

While WBC *BRCA1* methylation has been associated with an elevated hazard ratio of triple-negative breast cancer, to this end the quantitative contribution of normal tissue *BRCA1* methylation to TNBC and, potentially, non-TNBC, has remained unknown due to lack of studies evaluating concordant *BRCA1* methylation in tumor tissue and matched WBC. Analyzing *BRCA1* methylation in matched blood and tumor tissue of patients with both TNBC and non-TNBC, we found a strong correlation between tumor tissue and WBC *BRCA1* methylation in TNBC and tumors revealing a low ER expression (1–9%). In this group, 29.2% of tumors were *BRCA1*-methylated, and 19.4% harbored concordant tumor and WBC methylation. While no previous data exist regarding the incidence of concordant *BRCA1* methylation in tumor and WBCs, our findings for tumor methylation in total (somatic plus constitutional origin) aligns with the finding by Glodzik et al [5] of 30% of TNBCs carrying tissue *BRCA1* promoter methylation. Importantly, if a percentage as high as 20% of TNBCs/ER-low breast cancers having concordant WBC methylation is reproduced across other cohorts, this would mean that a substantially larger fraction of TNBCs may be caused by constitutional methylation than by pathogenic germline variants.

In contrast, among breast cancers with HER2 overexpression or ER levels of ≥10%, constitutional *BRCA1* tumor methylation was a rare event. However, interestingly, two out of three tumors with constitutional methylation revealed a basal-like gene expression signature.

Recently, we reported WBC *BRCA1* methylation to be predominantly monoallelic, enriched on the same allele across the vast majority of normal blood cells in affected adult individuals [24]. Here, we significantly extended this observation by showing concordant allele-specific methylation in normal and malignant cells in our cancer patients. This is consistent with a common clonal origin of methylated normal and cancer cells, which supports the hypothesis that these *BRCA1*-methylated tumors have arisen from methylated normal cells.

In addition, in four patients informative for methylation allele specificity in WBC, we found the tumor samples to reveal low-level *BRCA1* methylation, likely reflecting a fraction of benign cells in the biopsy. Here, we found similar allelic concordance for *BRCA1* methylation in WBC and the presumed normal breast tissue. While such low-level methylation in theory could reflect small subclones of somatically methylated tumor cells, the chance of allelic concordance across four paired datasets is <7%. In contrast, the finding of similar allele-specific methylation in WBC and normal breast tissue is what one would expect in cases of constitutional (prenatal) methylation. While the term “non-malignant cells” in a breast cancer biopsy covers different cell types [37] of ectodermal as well as mesenchymal origin, constitutional methylation generally affects cells derived from all three embryonic germ layers [13].

While our patient data are consistent with constitutional *BRCA1* methylation, a pivotal question is whether normal tissue *BRCA1* methylation develops as a complete promoter methylation (effectively repressing *BRCA1* expression) *in utero* or arise as a partially functional epimutation, subsequently developing into complete promoter methylation in normal cells postnatally. While we and others previously detected WBC *BRCA1* methylation among newborn girls [10, 44], these studies assessed methylation status by conventional analyses (qualitative methylation-specific PCR) preventing detailed assessment of allele specificity and quantitative characteristics of methylation. Notably, there is evidence showing that gene promoter methylation may develop in a step-wise manner [45], thus, methylation previously recorded by MSP in newborns could present a pre-methylation step or a qualitatively completely different methylation process from the one detected in normal cells of adults. In the present study, we found *BRCA1* methylation in newborn girls to qualitatively and quantitatively mirror the one seen in adult cancer patients and previously recorded in healthy adults [24], indicating that the methylation observed in newborns and in adults is the same molecular feature.

Taken together, these findings, in concert with our findings of similar allelic methylation status in WBC and tumor tissue in adults, are consistent with a common clonal origin of all *BRCA1*-methylated cells within each patient, indicating that methylation may have arisen as a single-cell event during early embryonic development with subsequent clonal expansion across all germ layers.

Analyzing WBCs collected from patients diagnosed with their breast cancer, it is important to exclude the possibility that WBC methylation is due to contamination from the tumor, either as circulating tumor DNA or circulating tumor cells. Yet, tumor contamination is unlikely to be the cause of WBC *BRCA1* methylation for several reasons. First; considering the number of circulating tumor cells, even among patients with a substantial cancer burden, such cells account for less than one in a million cells [46]. Second, as for circulating tumor DNA [47, 48], the plasma volume currently required for detection of tumor-derived genomic aberrations in blood samples is far above any possible plasma remnants in our WBC assay. Finally, in our recent study [24] we confirmed WBC *BRCA1* methylation in healthy women to predict subsequent incident TNBC as well as HGSOC >5 years after sampling, providing proof for normal cell *BRCA1* methylation to be a precursor for TNBC and HGSOC.

Cancer patients may have a different WBC subfraction composition as compared to healthy individuals. Thus, a potential uncertainty in the present study relates to cancer-related changes in the WBC subfraction composition. While global methylation patterns vary between leukocyte subfractions [49, 50], examining *BRCA1* methylation status across previously reported datasets from adults [51], newborns and corresponding 5-year old children [29, 52] we detected no difference in *BRCA1* methylation status between the different WBC subfractions [10]. Thus, the observed differences in *BRCA1* methylation may not be a consequence of differences in WBC subfraction composition.

Taken together, we consider these findings to validate and justify the use of WBC *BRCA1* methylation as a marker of constitutional methylation in most individuals, including patients diagnosed with their primary breast cancer.

In our recent case-control WHI study [24], WBC *BRCA1* methylation was associated with an increased risk of incident TNBC (hazard ratio, HR 2.5) and HGSOC (HR 1.8). Notably, in that study the median age at inclusion was 62 years, yet, TNBCs are known in general to be detected at an earlier age compared to other breast cancer subtypes [5]. Regarding HGSOC, the risk estimates in the WHI study is lower than that previously observed (HR 2.2–2.9) in a hospital-based cohort study in Norwegian women in which methylation was assessed by MSP [10]. Thus, the possibility exists that the lifetime risk for TNBC is higher than what we recorded in the WHI study. While the number of cases in the present study is limited, our finding that constitutional *BRCA1* methylation may account for 19.4% of all triple-negative and ER-low breast cancers is high given the observed *BRCA1* methylation frequency in the population (5.6% among non-cancer females in the US [24] and 9.0% among healthy newborns in the current study). Since constitutional *BRCA1* methylation affects a small fraction of normal cells in the individual, one may assume that the background incidence of *BRCA1* unmethylated breast cancers (including TNBC) is similar among carriers and non-carriers of *BRCA1* constitutional methylation. Based on this, our present data indicates that constitutional *BRCA1* methylation could be associated with a hazard ratio perhaps as high as 4–5 for TNBC/ER-low BC development [25].

In summary, we find constitutional *BRCA1* methylation, as defined by WBC methylation, to be linked to TNBC/ER-low BC. As for patients diagnosed with TNBC or ER-low tumors, our findings indicate that *BRCA1* promoter methylation should be explored as a potential risk factor for subsequent cancer development. Moreover, comparing cancers carrying the methylation on a constitutional versus somatic background should be performed to fully elucidate potential pathogenic consequences.The presence of *BRCA1* methylation on the same *BRCA1* allele in WBC and breast cancer DNA in the same patients adds strong support to the hypothesis that *BRCA1*-methylated tumors may arise from constitutionally *BRCA1-*methylated normal cells, likely initiated as an early, prenatal event [13]. Thus, our findings conceptually differ from normal tissue global methylation signatures designed for early cancer detection [53–55]. Our findings urge for further research exploring potential causes of *BRCA1* methylation, as well as studies exploring potential constitutional methylation of other tumor suppressor genes, with respect to cancer risk. To this end, cancers have generally been classified in two main groups; those arising on a background of pathogenic germline variants and those regarded as spontaneous tumors, with a grey zone of low-risk variants and genes, in-between. Our findings point toward the dawn of a new era, suggesting a substantial number of cancers may develop from early epimutated cell clones.

## Supplementary Information

The online version contains supplementary material available at …………..

**Additional file 1.** Contains Supplementary Methods, Supplementary Fig.s S1–S6 and Supplementary Table S1. **Fig. S1**: Genomic structure of the *BRCA1* promoter area, positions of CpGs, single-nucleotide variations, and amplified regions **Fig. S2**: Distributions of VEF values (CpG14–34 average methylation metric), corresponding density functions and cutoff bounds. **Fig. S3**: Scatter plots and density histograms showing the relation of VEF and beta values for blood and tumor samples. **Fig. S4**: Scatter plots and density histograms showing the relation of VEF and beta values for newborn blood samples. **Fig. S5**: PCR bias assessed for VEF and methylation beta values. **Fig. S6.** Intrinsic breast cancer subtypes based on gene expression analysis of tumors. **Table S1.** *BRCA1* methylation status in TNBC cases with pathogenic (germline or somatic) *BRCA1* mutations

## Data Availability

Haukeland University Hospital and the University of Bergen support the dissemination of research data that has been generated, and increased cooperation between investigators. Trial data is collected, stored, and disseminated according to institutional guidelines and in accordance with national laws and regulations to ensure the quality, integrity, and use of clinical data. Study protocol, including plan for statistical analyses, is available online via previous publications. Signed informed consent forms are stored at each participating hospital and are available for monitoring by regulatory authorities. After publication and upon formal request, raw data, including de-identified individual participant data and a data dictionary defining each field in the data set, will be shared according to institutional procedures. Requests are via a standard pro forma describing the nature of the proposed research and extent of data requirements. Data recipients are required to enter a formal data sharing agreement which describes the conditions for release and requirements for data transfer, storage, archiving, publication, and intellectual property. Requests are reviewed by the EPITAX, DDP and PETREMAC study teams in terms of scientific merit and ethical considerations, including patient consent. Data sharing is permitted if proposed projects have a sound scientific or patient benefit rationale, as agreed by the study team and with approval from the trials' co-investigators as required.
Samples from the MoBa study were analyzed blinded to the identity of the participants. Since BRCA1 WBC methylation has been found associated with an elevated risk of HGSOC and TNBC, under Norwegian law, this means that WBC BRCA1 methylation status may be considered predictive testing. Thus, our Regional Ethics Committee approved this part of the study provided no identification key or link to clinical information was stored. Thus, all molecular data from newborns in the present study were handled anonymously after analysis. Raw molecular data are available from the authors upon reasonable individual request, but any link to parameters in the MoBa databank is not possible.

## Acknowledgements

We gratefully acknowledge the technical assistance of N.K. Duong, C. Eriksen, E. De Faveri, and G. Iversen, University of Bergen and Haukeland University Hospital.

## Authors contribution

PEL and SK conceived the study. ON and SK developed the laboratory methods, and ON was in primary charge of the statistical analysis and performed all the epigenome analysis. PEL (EPITAX, DDP and PETREMAC) and HPE (DDP and PETREMAC) were in charge of the clinical studies. BG, SL, ESB, HE; JG and SG provided clinical material for the analysis. EAMJ was in charge of the immunochemistry, while SY, LM, BL and RL performed the epigenetic and subtype analysis. PEL, ON and SK drafted the manuscript, while all authors participated in editing of the final version.

## Data availability and materials

Haukeland University Hospital and the University of Bergen support the dissemination of research data that has been generated, and increased cooperation between investigators. Trial data is collected, stored, and disseminated according to institutional guidelines and in accordance with national laws and regulations to ensure the quality, integrity, and use of clinical data. Study protocol, including plan for statistical analyses, is available online via previous publications. Signed informed consent forms are stored at each participating hospital and are available for monitoring by regulatory authorities. After publication and upon formal request, raw data, including de-identified individual participant data and a data dictionary defining each field in the data set, will be shared according to institutional procedures. Requests are via a standard pro forma describing the nature of the proposed research and extent of data requirements. Data recipients are required to enter a formal data sharing agreement which describes the conditions for release and requirements for data transfer, storage, archiving, publication, and intellectual property. Requests are reviewed by the EPITAX, DDP and PETREMAC study teams in terms of scientific merit and ethical considerations, including patient consent. Data sharing is permitted if proposed projects have a sound scientific or patient benefit rationale, as agreed by the study team and with approval from the trials’ co-investigators as required.

Samples from the MoBa study were analyzed blinded to the identity of the participants. Since *BRCA1* WBC methylation has been found associated with an elevated risk of HGSOC and TNBC [24], under Norwegian law, this means that WBC *BRCA1* methylation status may be considered predictive testing. Thus, our Regional Ethics Committee approved this part of the study provided no identification key or link to clinical information was stored. Thus, all molecular data from newborns in the present study were handled anonymously after analysis. Raw molecular data are available from the authors upon reasonable individual request, but any link to parameters in the MoBa databank is not possible.

## Declarations

### Ethics approval and consent to participate

All participants in the clinical studies provided written informed consent, and the current study is covered by each consent. All studies were approved by the Regional Ethics Committee (reference numbers: 273/96-82.96, 06/3077 and 2015/1493).

### Competing interests

**Research Funding (***to Institution*): Astellas Oncology (BG), AstraZeneca (HPE, BG, SK, PEL, ESB), Celgene (BG), Novartis (HPE, PEL), Pfizer (HPE, SK, BG, PEL).

**Honoraria:** Amgen (HPE), AstraZeneca (HPE, BG, TA, EAMJ, SK, PEL), Abbvie (PEL), Bristol-Myers-Squibb (HPE, JG), Dagens Medisin (HPE, PEL), Eli Lilly (JG), HAI Interaktiv AS (HPE), MSD (JG), Novartis (HPE, JG, SK), Pfizer (HPE, EAJ, SK), Pierre Fabre (HPE, JG, SK, PEL), Roche (HPE, BG, TA, PEL).

**Consulting or Advisory Role:** Aptitude Health (HPE), Astellas Oncology (BG), AstraZeneca (ESB, JG, PEL), Daiichi Sankyo (ESB, HPE), Eli Lilly (ESB, HPE, JG), Gilead (ESB), Laboratorios Farmaceuticos Rovi (PEL), MSD (ESB, HPE, JG), Novartis (ESB, HPE, JG), Pfizer (HPE), Pierre Fabre (HPE), Roche (HPE, BG)

Expert Testimony: Pfizer (HPE).

**Travel, Accomodations, Expenses:** AstraZeneca (HPE), Pierre Fabre (HPE, BG, PEL), Roche (BG).

**Speakers’ Bureau:** Akademikonferens (PEL), Aptitude Health (PEL), AstraZeneca (JG), Bristol-Myers-Squibb (JG), MSD (JG), Novartis (JG), Pfizer (JG), Pierre Fabre (JG).

**Patents, Royalties, Other Intellectual Property:** Patent EP2389450 A1 (SK), Patent WO 2012/ 010661 (SK), Cytovation (PEL).

All remaining authors have declared no conflicts of interest.

### Author details

^1^K.G. Jebsen Center for Genome-Directed Cancer Therapy, Department of Clinical Science, University of Bergen; Bergen, Norway.

^2^Department of Oncology, Haukeland University Hospital; Bergen, Norway.

^3^Department of Hematology and Oncology, Stavanger University Hospital; Stavanger, Norway.

^4^Cancer Clinic, St. Olavs Hospital, Trondheim University Hospital; Trondheim, Norway.

^5^Department of Clinical and Molecular Medicine, Norwegian University of Science and Technology; Trondheim, Norway.

^6^Department of Oncology, University Hospital of North Norway; Tromsø, Norway.

^7^Department of Surgery, Haugesund Hospital; Haugesund, Norway.

^8^Department of Oncology, Akershus University Hospital; Lørenskog, Norway.

^9^Institute of Clinical Medicine, University of Oslo, Norway.

^10^Department of Pathology, Stavanger University Hospital; Stavanger, Norway.

^11^Department of Chemistry, Bioscience and Environmental Engineering; Stavanger University, Stavanger, Norway.

## REFERENCES

1. Esteller M: Epigenetics in cancer. N Engl J Med 2008, 358:1148–1159.

2. Li X, Yao X, Wang Y, Hu F, Wang F, Jiang L, Liu Y, Wang D, Sun G, Zhao Y: MLH1 promoter methylation frequency in colorectal cancer patients and related clinicopathological and molecular features. PLoS One 2013, 8:e59064.

3. Walsh T, Gulsuner S, Lee MK, Troester MA, Olshan AF, Earp HS, Perou CM, King MC: Inherited predisposition to breast cancer in the Carolina Breast Cancer Study. Npj Breast Cancer 2021, 7.

4. Kontorovich T, Cohen Y, Nir U, Friedman E: Promoter methylation patterns of ATM, ATR, BRCA1, BRCA2 and P53 as putative cancer risk modifiers in Jewish BRCA1/BRCA2 mutation carriers. Breast Cancer Research and Treatment 2009, 116:195–200.

5. Glodzik D, Bosch A, Hartman J, Aine M, Vallon-Christersson J, Reutersward C, Karlsson A, Mitra S, Nimeus E, Holm K, et al: Comprehensive molecular comparison of BRCA1 hypermethylated and BRCA1 mutated triple negative breast cancers. Nature Communications 2020, 11.

6. Eikesdal HP, Yndestad S, Elzawahry A, Llop-Guevara A, Gilje B, Blix ES, Espelid H, Lundgren S, Geisler J, Vagstad G, et al: Olaparib monotherapy as primary treatment in unselected triple negative breast cancer. Annals of Oncology 2021, 32:240–249.

7. Cunningham JM, Cicek MS, Larson NB, Davila J, Wang C, Larson MC, Song H, Dicks EM, Harrington P, Wick M, et al: Clinical Characteristics of Ovarian Cancer Classified by BRCA1, BRCA2, and RAD51C Status. Scientific Reports 2014, 4; art number 4026::1–7.

8. Rice JC, Futscher BW: Transcriptional repression of BRCA1 by aberrant cytosine methylation, histone hypoacetylation and chromatin condensation of the BRCA1 promoter. Nucleic Acids Research 2000, 28:3233–3239.

9. Joosse SA, Brandwijk KIM, Mulder L, Wesseling J, Hannemann J, Nederlof PM: Genomic Signature of BRCA1 Deficiency in Sporadic Basal-Like Breast Tumors. Genes Chromosomes & Cancer 2011, 50:71–81.

10. Lønning PE, Berge EO, Bjornslett M, Minsaas L, Chrisanthar R, Hoberg-Vetti H, Dulary C, Busato F, Bjorneklett S, Eriksen C, et al: White Blood Cell BRCA1 Promoter Methylation Status and Ovarian Cancer Risk. Annals of Internal Medicine 2018, 168:326-+.

11. Menghi F, Banda K, Kumar P, Straub R, Dobrolecki L, Rodriguez IV, Yost SE, Chandok H, Radke MR, Somlo G, et al: Genomic and epigenomic BRCA alterations predict adaptive resistance and response to platinum-based therapy in patients with triple-negative breast and ovarian carcinomas. Science Translational Medicine 2022, 14.

12. Tutt A, Tovey H, Cheang MCU, Kernaghan S, Kilburn L, Gazinska P, Owen J, Abraham J, Barrett S, Barrett-Lee P, et al: Carboplatin in BRCA1/2-mutated and triple-negative breast cancer BRCAness subgroups: the TNT Trial. Nature Medicine 2018, 24:628-+.

13. Lønning PE, Eikesdal HP, Loes IM, Knappskog S: Constitutional Mosaic Epimutations – a hidden cause of cancer? Cell Stress 2019, 3:118–135.

14. Evans DGR, van Veen EM, Byers HJ, Wallace AJ, Ellingford JM, Beaman G, Santoyo-Lopez J, Aitman TJ, Eccles DM, Lalloo FI, et al: A Dominantly Inherited 5’ UTR Variant Causing Methylation-Associated Silencing of BRCA1 as a Cause of Breast and Ovarian Cancer. American Journal of Human Genetics 2018, 103:213–220.

15. Laner A, Benet-Pages A, Neitzel B, Holinski-Feder E: Analysis of 3297 individuals suggests that the pathogenic germline 5’-UTR variant BRCA1 c.-107A > T is not common in south-east Germany. Familial Cancer 2020, 19:211–213.

16. de Jong VMT, Pruntel R, Steenbruggen TG, Bleeker FE, Nederlof P, Hogervorst FBL, Linn SC: Identifying the BRCA1 c.-107A > T variant in Dutch patients with a tumor BRCA1 promoter hypermethylation. Familial Cancer 2022: DOI: 10.1007/s10689-10022-00314-z.

17. Prajzendanc K, Domagala P, Hybiak J, Rys J, Huzarski T, Szwiec M, Tomiczek-Szwiec J, Redelbach W, Sejda A, Gronwald J, et al: BRCA1 promoter methylation in peripheral blood is associated with the risk of triple-negative breast cancer. International Journal of Cancer 2020, 146:1293–1298.

18. Snell C, Krypuy M, Wong EM, Loughrey MB, Dobrovic A: BRCA1 promoter methylation in peripheral blood DNA of mutation negative familial breast cancer patients with a BRCA1 tumour phenotype. Breast Cancer Research 2008, 10: R12; 10.1186/bcr1858:1-8.

19. Wong EM, Southey MC, Fox SB, Brown MA, Dowty JG, Jenkins MA, Giles GG, Hopper JL, Dobrovic A: Constitutional Methylation of the BRCA1 Promoter Is Specifically Associated with BRCA1 Mutation-Associated Pathology in Early-Onset Breast Cancer. Cancer Prevention Research 2011, 4:23–33.

20. Hansmann T, Pliushch G, Leubner M, Kroll P, Endt D, Gehrig A, Preisler-Adams S, Wieacker P, Haaf T: Constitutive promoter methylation of BRCA1 and RAD51C in patients with familial ovarian cancer and early-onset sporadic breast cancer. Human Molecular Genetics 2012, 21:4669–4679.

21. Iwamoto T, Yamamoto N, Taguchi T, Tamaki Y, Noguchi S: BRCA1 promoter methylation in peripheral blood cells is associated with increased risk of breast cancer with BRCA1 promoter methylation. Breast Cancer Res Treat 2011, 129:69–77.

22. Bosviel R, Michard E, Lavediaux G, Kwiatkowski F, Bignon YJ, Bernard-Gallon DJ: Peripheral blood DNA methylation detected in the BRCA1 or BRCA2 promoter for sporadic ovarian cancer patients and controls. Clinica Chimica Acta 2011, 412:1472–1475.

23. Azzollini J, Pesenti C, Pizzamiglio S, Fontana L, Guarino C, Peissel B, Plebani M, Tabano S, Sirchia SM, Colapietro P, et al: Constitutive BRCA1 Promoter Hypermethylation Can Be a Predisposing Event in Isolated Early-Onset Breast Cancer. Cancers 2019, 11.

24. Lønning PE, Nikolaienko O, Pan K, Kurian AW, Eikesdal HP, Pettinger M, Anderson GL, Prentice RL, Chlebowski RT, Knappskog S: Constitutional BRCA1 Methylation and Risk of Incident Triple-Negative Breast Cancer and High-grade Serous Ovarian Cancer. Jama Oncology 2022, 8:1579–1587.

25 Bruzzi P, Green SB, Byar DP, Brinton LA, Schairer C: ESTIMATING THE POPULATION ATTRIBUTABLE RISK FOR MULTIPLE RISK-FACTORS USING CASE-CONTROLDATA. American Journal of Epidemiology 1985, 122:904–913.

26. Chrisanthar R, Knappskog S, Lokkevik E, Anker G, Ostenstad B, Lundgren S, Risberg T, Mjaaland I, Skjonsberg G, Aas T, et al: Predictive and prognostic impact of TP53 mutations and MDM2 promoter genotype in primary breast cancer patients treated with epirubicin or paclitaxel. PLos One 2011, 6:e19249.

27. Venizelos A, Engebrethsen C, Deng W, Geisler J, Geisler S, Iversen GT, Aas T, Aase HS, Seyedzadeh M, Steinskog ES, et al: Clonal evolution in primary breast cancers under sequential epirubicin and docetaxel monotherapy. Genome Medicine 2022, 14.

28. Magnus P, Birke C, Vejrup K, Haugan A, Alsaker E, Daltveit AK, Handal M, Haugen M, Hoiseth G, Knudsen GP, et al: Cohort Profile Update: The Norwegian Mother and Child Cohort Study (MoBa). International Journal of Epidemiology 2016, 45:382–388.

29. Gervin K, Page CM, Aass HC, Jansen MA, Fjeldstad HE, Andreassen BK, Duijts L, van Meurs JB, van Zelm MC, Jaddoe VW, et al: Cell type specific DNA methylation in cord blood: A 450K-reference data set and cell count-based validation of estimated cell type composition. Epigenetics 2016, 11:690–698.

30. Perou CM, Sorlie T, Eisen MB, van de Rijn M, Jeffrey SS, Rees CA, Pollack JR, Ross DT, Johnsen H, Akslen LA, et al: Molecular portraits of human breast tumours. Nature 2000, 406:747–752.

31. Yates LR, Gerstung M, Knappskog S, Desmedt C, Gundem G, Van Loo P, Aas T, Alexandrov LB, Larsimont D, Davies H, et al: Subclonal diversification of primary breast cancer revealed by multiregion sequencing. Nature Medicine 2015, 21:751-+.

32. Landrum MJ, Chitipiralla S, Brown GR, Chen C, Gu BS, Hart J, Hoffman D, Jang W, Kaur K, Liu CL, et al: ClinVar: improvements to accessing data. Nucleic Acids Research 2020, 48:D835–D844.

33. Nikolaienko O, Lønning PE, Knappskog S: epialleleR: an R/BioC package for sensitive allele-specific methylation analysis in NGS data. bioRxiv 2022:2022.2006.2030.498213.

34. Lonning PE, Nikolaienko O, Pan K, Kurian AW, Eikesdal HP, Pettinger M, Anderson GL, Prentice RL, Chlebowski RT, Knappskog S: Constitutional BRCA1 Methylation and Risk of Incident Triple-Negative Breast Cancer and High-grade Serous Ovarian Cancer. Jama Oncology 2022, 8:1579–1587.

35. Magnus P, Birke C, Vejrup K, Haugan A, Alsaker E, Daltveit AK, Handal M, Haugen M, Hoiseth G, Knudsen GP, et al: Cohort Profile Update: The Norwegian Mother and Child Cohort Study (MoBa). Int J Epidemiol 2016, 45:382–388.

36. Iwamoto T, Booser D, Valero V, Murray JL, Koenig K, Esteva FJ, Ueno NT, Zhang J, Shi WW, Qi Y, et al: Estrogen Receptor (ER) mRNA and ER-Related Gene Expression in Breast Cancers That Are 1% to 10% ER-Positive by Immunohistochemistry. J Clin Oncol 2012, 30:729–734.

37. Wagner J, Rapsomaniki MA, Chevrier S, Anzeneder T, Langwieder C, Dykgers A, Rees M, Ramaswamy A, Muenst S, Soysa SD, et al: A Single-Cell Atlas of the Tumor and Immune Ecosystem of Human Breast Cancer. Cell 2019, 177:1330-+.

38. Esteller M.: Molecular origins of cancer: Epigenetics in cancer. New England Journal of Medicine 2008, 358:1148–1159.

39. Hegi ME, Diserens A, Gorlia T, Hamou M, de Tribolet N, Weller M, Kros JM, Hainfellner JA, Mason W, Mariani L, et al: MGMT gene silencing and benefit from temozolomide in glioblastoma. New England Journal of Medicine 2005, 352:997–1003.

40. Della Monica R, Cuomo M, Buonaiuto M, Costabile D, Franca RA, De Caro MD, Catapano G, Chiariotti L, Visconti R: MGMT and Whole-Genome DNA Methylation Impacts on Diagnosis, Prognosis and Therapy of Glioblastoma Multiforme. International Journal of Molecular Sciences 2022, 23.

41. Feinberg AP, Tycko B: The history of cancer epigenetics. Nature Reviews Cancer 2004, 4:143–153.

42. Hitchins MP, Wong JJL, Suthers G, Suter CM, Martin DIK, Hawkins NJ, Ward RL: Brief report: Inheritance of a cancer-associated MLH1 germ-line epimutation. New England Journal of Medicine 2007, 356:697–705.

43. Hitchins MP: Constitutional epimutation as a mechanism for cancer causality and heritability? Nature Rev Cancer 2015, 15:181–194.

44. Al-Moghrabi N, Al-Showimi M, Al-Yousef N, Al-Shahrani B, Karakas B, Alghofaili L, Almubarak H, Madkhali S, Al Humaidan H: Methylation of BRCA1 and MGMT genes in white blood cells are transmitted from mothers to daughters. Clinical Epigenetics 2018, 10.

45. Skvortsova K, Masle-Farquhar E, Luu PL, Song JZ, Qu WJ, Zotenko E, Gould CM, Du Q, Peters TJ, Colino-Sanguino Y, et al: DNA Hypermethylation Encroachment at CpG Island Borders in Cancer Is Predisposed by H3K4 Monomethylation Patterns. Cancer Cell 2019, 35:297-+.

46. Rack B, Schindlbeck C, Andergassen U, Lorenz R, Zwingers T, Schneeweiss A, Lichtenegger W, Beckmann MW, Sommer HL, Pantel K, et al: Prognostic relevance of circulating tumor cells in the peripheral blood of primary breast cancer patients. Ca Res 2010, 70(Suppl):S6–5, 93s.

47. Liu MC, Oxnard GR, Klein EA, Swanton C, Seiden MV, Consortium C: Sensitive and specific multi-cancer detection and localization using methylation signatures in cell-free DNA. Annals of Oncology 2020, 31:745–759.

48. Li YZ, Fan ZY, Meng YF, Liu SJ, Zhan HX: Blood-based DNA methylation signatures in cancer: A systematic review. Biochimica Et Biophysica Acta-Molecular Basis of Disease 2023, 1869.

49. Houseman EA, Accomando WP, Koestler DC, Christensen BC, Marsit CJ, Nelson HH, Wiencke JK, Kelsey KT: DNA methylation arrays as surrogate measures of cell mixture distribution. Bmc Bioinformatics 2012, 13; art number 86:1–16.

50. Fridley BL, Armasu SM, Cicek MS, Larson MC, Wang C, Winham SJ, Kalli KR, Koestler DC, Rider DN, Shridhar V, et al: Methylation of leukocyte DNA and ovarian cancer: relationships with disease status and outcome. Bmc Medical Genomics 2014, 7; art no 21::1–12.

51. Reinius LE, Acevedo N, Joerink M, Pershagen G, Dahlen SE, Greco D, Soderhall C, Scheynius A, Kere J: Differential DNA Methylation in Purified Human Blood Cells: Implications for Cell Lineage and Studies on Disease Susceptibility. Plos One 2012, 7:1–13.

52. Martino DJ, Tulic MK, Gordon L, Hodder M, Richman T, Metcalfe J, Prescott SL, Saffery R: Evidence for age-related and individual-specific changes in DNA methylation profile of mononuclear cells during early immune development in humans. Epigenetics 2011, 6:1085–1094.

53. Widschwendter M, Jones A, Evans I, Reiser D, Dillner J, Sundstrom K, Steyerberg EW, Vergouwe Y, Wegwarth O, Rebitschek FG, et al: Epigenome-based cancer risk prediction: rationale, opportunities and challenges. Nature Reviews Clinical Oncology 2018, 15:292–309.

54. Barrett JE, Herzog C, Jones A, Leavy OC, Evans I, Knapp S, Reisel D, Nazarenko T, Kim Y-N, Franchi D, et al: The WID-BC-index identifies women with primary poor prognostic breast cancer based on DNA methylation in cervical samples. Nature Communications 2022, 13.

55. Barrett JE, Jones A, Evans I, Reisel D, Herzog C, Chindera K, Kristiansen M, Leavy OC, Manchanda R, Bjorge L, et al: The DNA methylome of cervical cells can predict the presence of ovarian cancer. Nature Communications 2022, 13.

