## Additional File 1 for "Prenatal *BRCA1* epimutations contribute significantly to triple-negative breast cancer development"

### Supplementary Information

#### *Nucleic acid isolation and control DNA*

For paired tumour and WBC samples, genomic DNA for methylation analyses was extracted using QIAamp DNA Mini kit (Qiagen, Valencia, CA) according to the manufacturer's instructions.

For cord blood samples from newborns, details on genomic DNA extraction and storage were described elsewhere.[1]

Total RNA for gene expression analysis was extracted from tumor tissue using the RNeasy Mini kit with an on-column DNase digestion according to the manufacturer's protocol (Qiagen, Valencia, CA). RNA quality was determined by UV absorption on a NanoDrop spectrophotometer and RNA concentration was determined with Qubit fluorometer (Thermo Fisher Scientific, Waltham, MA). RNA integrity numbers (RIN) were estimated using the RNA 6000 Nano assay (Agilent Technologies, Santa Clara, CA) run on the 2100 Bioanalyzer Instrument (Agilent Technologies, Santa Clara, CA). RIN values above 7 were required, but RIN >6 was accepted if no further material was available.

Human cell line HCT116 DKO (*DNMT1*$^{(-/-)}$ and *DNMT3B*$^{(-/-)}$) Non-Methylated and Methylated DNA control samples (Zymo Research, Irvine, CA; cat.no. D5014-1 and D5014-2 respectively) and their mixes with varying ratios were used to test methylation sequencing assay sensitivity.

#### *Molecular subtyping of tumors*

For molecular subtyping of samples from DDP and PETREMAC trials, RNA sequencing was applied. An input of 200–600 ng total RNA was converted to dual-indexed libraries using either TruSeq Stranded Human Total RNA Ribozero Gold Library Prep Kit (PETREMAC trial) or Illumina Stranded Total RNA Prep Ligation with Ribo-Zero plus kit (DDP trial), (Illumina, San Diego, CA). Library average sizes and quality were assessed using the DNA 1000 assay (Agilent Technologies, Santa Clara, CA) run on the 2100 Bioanalyzer. PETREMAC libraries were quantified by real-time PCR using KAPA Library Quantification kits (Kapa Biosystems, Wilmington, MA) with Lightcycler 480 II (Roche, Basel, Switzerland) while DDP libraries were quantified with Qubit dsDNA HS Assay Kit. Libraries were normalized, pooled and then sequenced on a NovaSeq 6000 (Illumina, San Diego, CA) using 2x100 cycles, providing a minimum of 70 million reads per sample. Base call files were processed using Illumina DRAGEN Bio-IT Platform v3.8.4. Samples were demultiplexed by the DRAGEN BCL converter, and gene expression data processed using the DRAGEN RNA pipeline mapping against GRCh38. Duplicate markings were enabled, but duplicate reads were not removed. An annotation file for ALT-aware mapping was downloaded from GENCODE (https://www.gencodegenes.org/human/).

A post hoc gene expression analysis was performed based on global RNA sequencing of pretreatment biopsies to assign the tumors to intrinsic breast cancer subtypes.[2] Briefly, genes with 0 counts were removed and raw expression data was normalized by variance stabilizing transformation using DESeq2.[3] Then, intrinsic subtypes were assigned using the R package Genefu v2.28.0,[4] using the centroids published by Parker et al.[5]

Molecular subtyping of samples from the EPITAX trial was performed based on previously described mRNA expression arrays.[6]

#### *BRCA1 methylation assay design*

The genomic structure of the *BRCA1* promoter region, location of CpG dinucleotides and four PCR amplicons are shown in the Supplementary Fig. S1. The genomic coordinates for the individual CpGs and entire amplicons, primer sequences and experimental conditions used for amplification as well as complete experimental details on library preparation, sequencing and data analysis were described previously.[7]

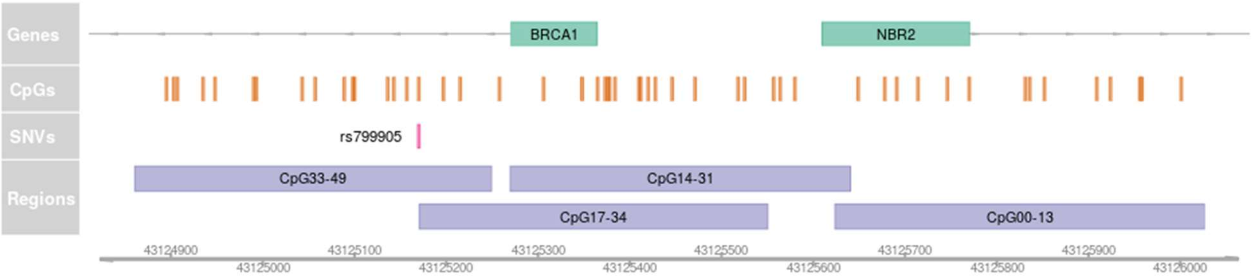

**Supplementary Figure S1.** Genomic structure of the *BRCA1* promoter area, positions of CpGs, single-nucleotide variations, and amplified regions.[7]

#### *Assay characteristics for paired blood and tumour samples*

Out of the four amplicons covering the *BRCA1* promoter region, amplicons CpG14–31 and CpG17–34 cover the region used as the main metric for *BRCA1* methylation in our previous work, in which DNA methylation was found to be associated with the risk of TNBC and HGSOC.[7] In the present analyses, the same region/metric was used; i.e. the average frequencies of hypermethylated epialleles covered by amplicons CpG14–31 and CpG17–34 were used to assess *BRCA1* promoter methylation (further referred to as region CpG14–34). Compared to genomic region-averaged beta values, this combined variant epiallele frequency (VEF) metric had lower minimum values as well as wider range of detected values (Supplementary Fig. S2 and S3).

Positivity cutoff value for *BRCA1* methylation in patient blood samples (equals 6.96e-04 for the region CpG14–34) was computationally defined as VEF value with the lowest probability in the range [ISR, MTS] (where ISR is index swap rate of 1.4e-04, and MTS—maximum theoretical sensitivity of 8.1e-04, as previously described in[7]; Supplementary Fig. S2A).

Positivity cutoff value for *BRCA1* methylation in tumor samples (equals 4.71e-02 for the region CpG14–34; Supplementary Fig. S2B) was computationally defined as VEF value with the lowest probability in the range [1e-02, 1e-01]. Here, the higher cutoff bounds were set in order to exclude samples with low-level mosaic *BRCA1* methylation in normal cells present in the biopsies, which should be expected in case of constitutional methylation affecting different tissues in the body. Thus, cutoffs were set to detect tumors with clonal expansions of *BRCA1* methylated cells.

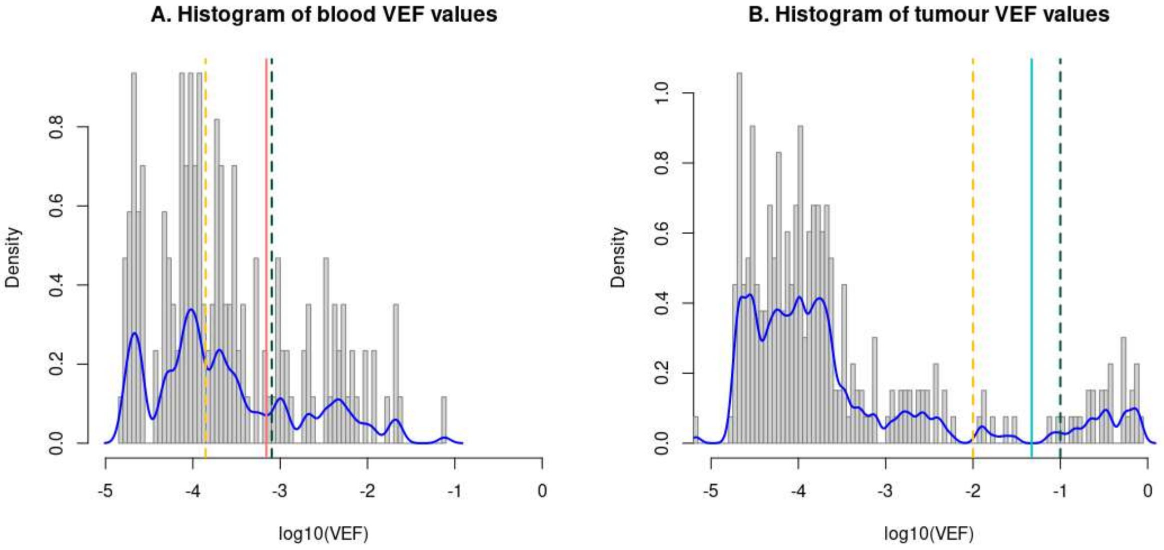

**Supplementary Figure S2.** Distributions of VEF values (CpG14–34 average methylation metric), corresponding density functions (blue lines) and cutoff bounds (dashed lines), positivity cutoffs for blood (light red line) and tumor (cyan line) samples.

Comparing the results of the present approach with our previously reported assessment of methylation for a subset of tumor samples[8] (*N*=32, TNBC cases) using methylation-specific quantitative PCR (MSP) revealed that samples with VEF of 4.55e-03 and higher were previously characterized as MSP-positive, while from 2.86e-03 and lower as MSP-negative (even though 200 ng of template DNA was used for MSP assay). This is in line with previously reported sensitivity of MSP assays of about 0.1% of methylated DNA[9] and confirms superior sensitivity of NGS-based approach and its validity in order to detect low-frequency methylation events.

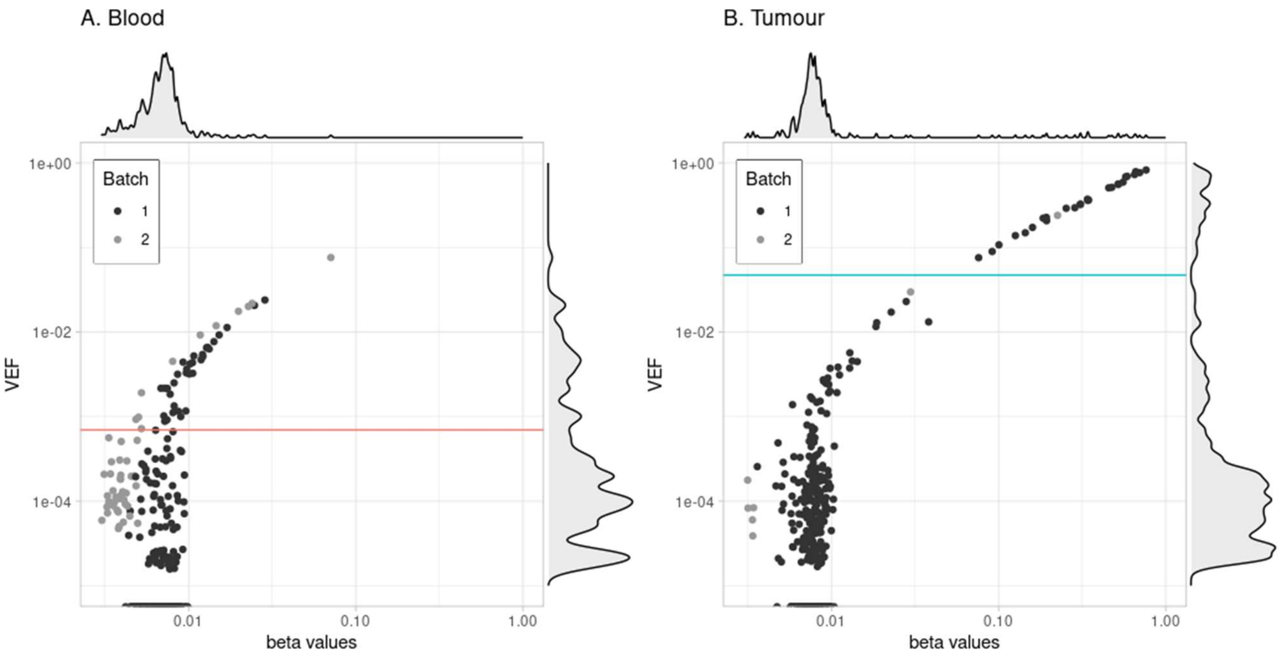

**Supplementary Figure S3.** Scatter plots and density histograms showing the relation of VEF and beta values for blood (A) and tumor (B) samples. Light red and cyan lines show corresponding methylation positivity cutoffs. Light grey and dark grey dots represent samples processed using two different batches of bisulfite conversion kits.

#### *Assay characteristics for newborn blood samples*

The same metric as described above (average VEF for amplicons CpG14–31 and CpG17–34) was used to characterize methylation in the blood samples from newborns. The same cutoff for methylation positivity as defined above for patient blood samples (equals 6.96e-04) was used to categorize newborn blood samples. Similarly to the characteristics described above, VEF metric had lower minimum values, wider range of detected values, and was less affected by batch effects, as compared to genomic region-averaged beta values (Supplementary Fig. S4).

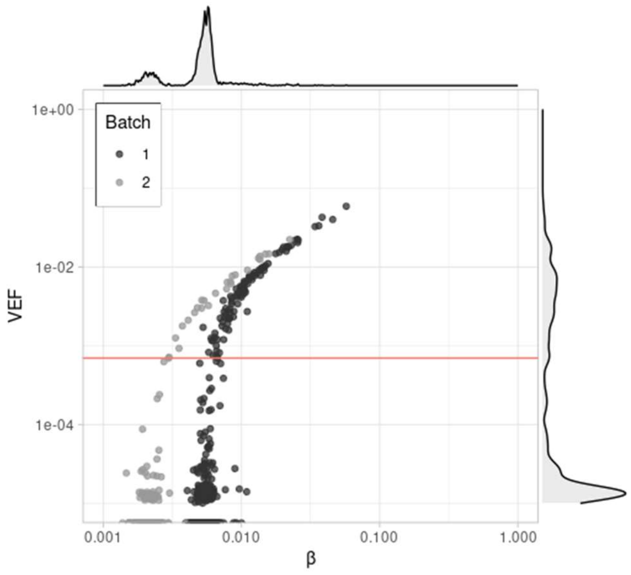

**Supplementary Figure S4.** Scatter plots and density histograms showing the relation of VEF and beta values for newborn blood samples. Light red line shows methylation positivity cutoff defined for patient blood samples above. Light gray and dark gray dots represent samples processed using two different batches of bisulfite conversion kits.

#### *Level of PCR bias*

Methylation control samples showed moderate preference for amplification of hypomethylated alleles (PCR bias), although theoretical and observed VEF values for hypermethylated alleles were highly concordant (b=0.6695, standard error of 0.022, as described in[10,11], versus beta values' b of 0.6069, standard error of 0.068; Supplementary Fig. S5). No PCR bias correction was performed prior the analysis.

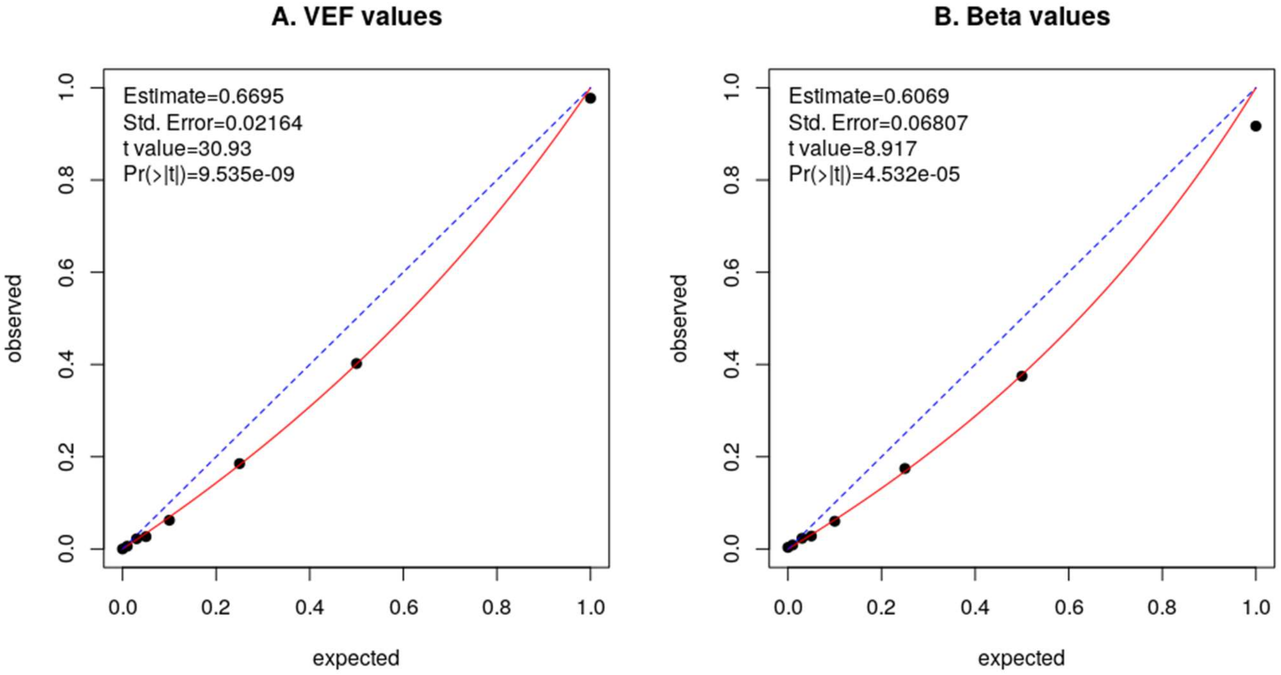

**Supplementary Figure S5.** PCR bias assessed for VEF and methylation beta values.

##### *BRCA1 methylation in subtypes of breast cancer*

TNBC and HER2– / ER<10% tumors were predominantly basal-like. Notably, among HER2–/ER>10% and HER2+ tumors four out of seven with *BRCA1* methylation in tumor or both tumor and blood were also basal-like (Supplementary Fig. S6).

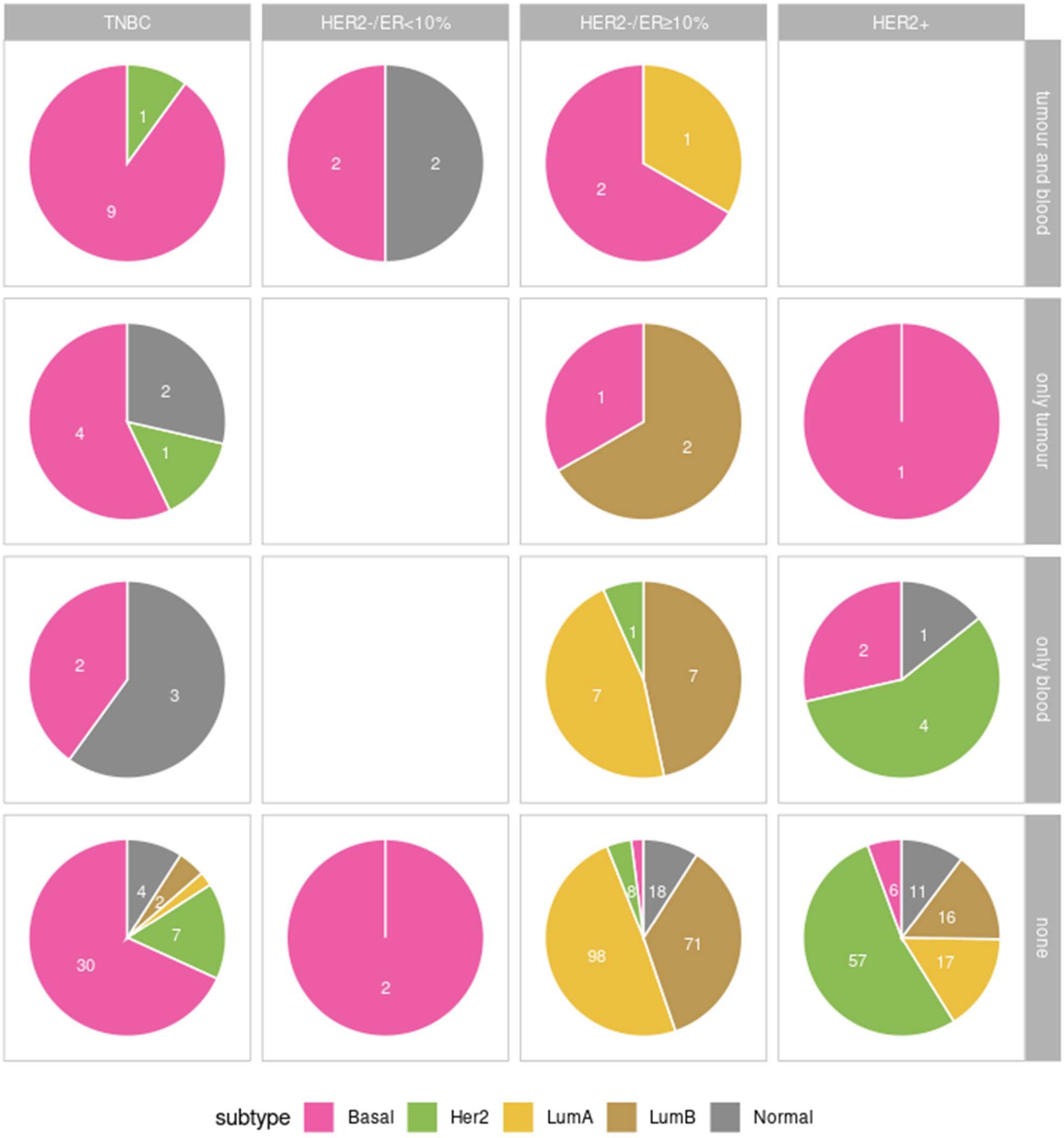

**Supplementary Figure S6.** Intrinsic breast cancer subtypes based on gene expression analysis of tumors. Pie charts are split by receptor expression status (columns) and *BRCA1* methylation status in blood and in tumor (rows). Basal: basal-like, HER2: HER2 enriched, lumA: luminal A, lumB: luminal B and Normal: normal-like subtypes.

#### *Distribution of BRCA1 pathogenic variants*

No association was observed between *BRCA1* methylation and *BRCA1* pathogenic variants (neither somatic nor germline variants; Supplementary Table 1.)

**Supplementary Table S1.** *BRCA1* methylation status among *N*=9 TNBC cases with pathogenic (germline or somatic) *BRCA1* mutations

| | | Methylated blood | Unmethylated blood |
|---|---|---|---|
| *BRCA1* germline variant | Methylated tumor | 0 | 0 |
| | Unmethylated tumor | 0 | 5 |
| *BRCA1* somatic mutation | Methylated tumor | 0 | 1 |
| | Unmethylated tumor | 0 | 3 |

#### References

1. Paltiel L, Anita H, Skjerden T, Harbak K, Bækken S, Kristin SN, et al. The biobank of the Norwegian Mother and Child Cohort Study – present status. Norsk Epidemiologi [Internet]. 2014 [cited 2023 Feb 1];24. Available from: https://www.ntnu.no/ojs/index.php/norepid/article/view/1755

2. Perou CM, Sørlie T, Eisen MB, van de Rijn M, Jeffrey SS, Rees CA, et al. Molecular portraits of human breast tumours. Nature. 2000;406:747–52.

3. Love MI, Huber W, Anders S. Moderated estimation of fold change and dispersion for RNA-seq data with DESeq2. Genome Biology. 2014;15:550.

4. Haibe-Kains B, Desmedt C, Loi S, Culhane AC, Bontempi G, Quackenbush J, et al. A Three-Gene Model to Robustly Identify Breast Cancer Molecular Subtypes. JNCI: Journal of the National Cancer Institute. 2012;104:311–25.

5. Parker JS, Mullins M, Cheang MCU, Leung S, Voduc D, Vickery T, et al. Supervised Risk Predictor of Breast Cancer Based on Intrinsic Subtypes. JCO. Wolters Kluwer; 2009;27:1160–7.

6. Poduval DB, Ognedal E, Sichmanova Z, Valen E, Iversen GT, Minsaas L, et al. Assessment of tumor suppressor promoter methylation in healthy individuals. Clin Epigenetics. 2020;12:131.

7. Lønning PE, Nikolaienko O, Pan K, Kurian AW, Eikesdal HP, Pettinger M, et al. Constitutional BRCA1 Methylation and Risk of Incident Triple-Negative Breast Cancer and High-grade Serous Ovarian Cancer. JAMA Oncol. 2022;8:1579–87.

8. Eikesdal HP, Yndestad S, Elzawahry A, Llop-Guevara A, Gilje B, Blix ES, et al. Olaparib monotherapy as primary treatment in unselected triple negative breast cancer☆. Annals of Oncology. 2021;32:240–9.

9. Herman JG, Graff JR, Myöhänen S, Nelkin BD, Baylin SB. Methylation-specific PCR: a novel PCR assay for methylation status of CpG islands. PNAS. National Academy of Sciences; 1996;93:9821–6.

10. Warnecke PM, Stirzaker C, Melki JR, Millar DS, Paul CL, Clark SJ. Detection and measurement of PCR bias in quantitative methylation analysis of bisulphite-treated DNA. Nucleic Acids Research. 1997;25:4422–6.

11. Moskalev EA, Zavgorodnij MG, Majorova SP, Vorobjev IA, Jandaghi P, Bure IV, et al. Correction of PCR-bias in quantitative DNA methylation studies by means of cubic polynomial regression. Nucleic Acids Research. 2011;39:e77–e77.
